# COVID-19 testing and vaccine willingness: Cross-sectional survey in a culturally diverse community in Sydney, Australia

**DOI:** 10.1101/2021.10.25.21265503

**Authors:** Julie Ayre, Danielle M Muscat, Olivia Mac, Carys Batcup, Erin Cvejic, Kristen Pickles, Hankiz Dolan, Carissa Bonner, Dana Mouwad, Dipti Zachariah, Una Turalic, Yyvonne Santalucia, Tingting Chen, Gordana Vasic, Kirsten McCaffery

## Abstract

**Objective:** The current study examined patterns in COVID-19 testing and vaccination intentions across multiple language groups in Greater Western Sydney, Australia.

**Methods:** Participants completed a cross-sectional survey available from March 21 to July 9, 2021 in Sydney, Australia. Surveys were available in English or translated (11 languages). Participants could complete surveys independently or with support from bilingual staff. Logistic regression models using post-stratification weighted frequencies identified factors associated with testing and vaccination intentions.

**Results:** Most of the 708 participants (88%, n=622) were not born in Australia; 31% reported that they did not speak English well or at all (n=220); 70% had no tertiary qualifications (n=497); and 41% had inadequate health literacy (n=290). Most participants reported high testing intention (77.2%, n=546), with differences observed across language groups (p<0.001). The most frequently reported barrier to testing was concerns about infection at the clinic (26.1%). Half (53.0%) reported willingness to get a COVID-19 vaccine if recommended to them (n=375); 18% were unwilling (n=127), and the remainder unsure (29%, n=205). These proportions varied significantly by language group (p<0.001). Participants were more likely to be unwilling/hesitant if they were female (p=0.02) or did not use Australian commercial information sources (p=0.01). Concerns about side effects (30.4%, n=102) and safety (23.9%, n=80), were key reported barriers to vaccination.

**Conclusion:** Different language groups have unique and specific needs to support uptake of COVID-19 testing and vaccination. Health services must work collaboratively with culturally and linguistically diverse communities to provide tailored support to encourage COVID-19 testing and vaccination.

**Lay summary:**

- 708 adults living in Sydney, Australia, who did not speak English as their main language at home took part in a survey about COVID-19 vaccination and testing.
- Participants could complete the survey online (English/translated) or with support from bilingual staff. The survey was available in 11 languages.
- Three quarters of participants (77%) reported they would get tested for COVID-19 if they had symptoms ‘no matter what.’ The most common barrier was concern about getting infected at the testing clinic.
- 53% of participants reported that they would get a COVID-19 vaccine if it was recommended to them. 18% reported that they wouldn’t get the vaccine, and 29% were unsure. The main barriers were concerns about vaccine side effects and safety.
- Intentions to get tested for COVID-19 or to get vaccinated varied significantly across language groups. Participants who were female, or who did not use Australian commercial information sources were more likely to be unwilling or unsure about getting a COVID-19 vaccine.
- Different language groups have unique and specific needs to support uptake of COVID-19 testing and vaccination. Health services must work collaboratively with culturally and linguistically diverse communities to provide tailored support to encourage COVID-19 testing and vaccination.

---

In 2021, vaccination became the main COVID-19 management strategy to enable countries to ease restrictions, open up the economy, and protect those who are most vulnerable from serious consequences of the virus(Australian Government, 2021b). In Australia COVID-19 vaccine rollout began in February 2021 and is underway at the time of writing. During the rollout, testing for COVID-19 symptoms (e.g. fever, cough, sore throat) continues to be another important management strategy. Testing and subsequent contact tracing of positive cases provides essential data about where, when, and how the virus is spreading, and informs decisions about how to contain outbreaks (e.g. through short-term localised restriction of movement).

In Australia, at the time of writing, COVID-19 vaccines were not mandatory. Australia has taken a state-based and phased approach to rollout, with eligibility focusing initially on frontline healthcare workers, older people, Aboriginal and Torres Strait Islander people, and people with specified pre-existing underlying medical conditions(Australian Government Department of Health, 2020). Two vaccines were available at the time of the study: Pfizer (Comirnaty) and AstraZeneca (Vaxzevria), although the supply of Pfizer was much more limited. Though both are widely used and accepted internationally in countries such as the UK, the AstraZeneca vaccine has drawn media attention due to a very small increased risk of thrombosis with thrombocytopenia (a side effect that can have serious consequences), and reports of poorer vaccine effectiveness compared to Pfizer(Picheta, 2021). This, coupled with the Australian Technical Advisory Group on Immunisation (ATAGI)’s changing advice about Pfizer as the ‘preferred vaccine’ for certain age groups likely reduced community willingness to get the AstraZeneca vaccine in the first half of 2021, which slowed the rollout because it was the only option available to many community members at that time(Leask et al., 2021).

Australian nationally representative surveys suggest that in the lead up to the vaccine rollout in February 2021, 73% of Australians agreed that they would get a COVID-19 vaccination when it is available and recommended to them(Australian Bureau of Statistics, 2021a). By April and May this was 68%, returning to 73% in June(Australian Bureau of Statistics, 2021a, Australian Bureau of Statistics, 2021b). Our Australia-wide surveys in April and November 2020, before vaccines became available, suggested that main barriers to COVID-19 vaccination included concerns about the vaccines’ safety, distrust of the government or the vaccine, and needing more information(Dodd et al., 2021, Dodd et al., 2020a, Dodd et al., 2020b). No differences in vaccine intention were observed for people who spoke a language other than English at home, though people with lower health literacy and less education had lower vaccine intentions. This finding is consistent with another nationally representative survey (sample recruited August 2020)(Edwards et al., 2021). However, it should be noted that these surveys did not adequately engage culturally and linguistically diverse communities in Australia, who were underrepresented in these samples. As such, there is only limited Australian research about COVID-19 vaccine intentions and barriers in these communities.

Since April 24^th^ 2020, all Australians have been encouraged to get tested for COVID-19 if they experience even mild symptoms. However nationally representative surveys estimate that the proportion of Australians reporting that they would definitely get a COVID-19 test if they had mild respiratory infection symptoms is relatively low, ranging between 45% and 51% between December 2020 and June 2021(Australian Bureau of Statistics, 2021a, Australian Bureau of Statistics, 2021b). Our survey in November 2020 highlighted that the most common barriers to testing included having symptoms but not believing it was COVID (e.g. hayfever) (15%), preferring to self-isolate (13%), feeling that symptoms were not severe enough (11%), and concern that the test was painful (10%)(Bonner et al., 2021).

The above research provides useful data on COVID-19 attitudes and intentions in general Australian samples. However, only a small proportion of participants had inadequate health literacy or spoke a language other than English at home. This limits our capacity to understand the needs of Australian communities that are typically understudied and underserved, such as those that are culturally and linguistically diverse. This is critically important; in other countries such as the UK and the US, research indicates that COVID-19 testing rates and vaccine acceptability are often lower for people in culturally and linguistically diverse communities(Razai et al., 2021, Khubchandani and Macias, 2021), with calls to prioritise testing and vaccination efforts for these groups(Dodds and Fakoya, 2020, Al-Oraibi et al., 2021).

The current study aimed to fill this research gap by describing patterns in COVID-19 testing intentions and vaccine willingness in a culturally and linguistically diverse sample in Sydney, Australia, during the period March 21 to July 9, 2021; and to identify key barriers to COVID-19 testing and vaccine willingness within these groups.

## Methods

### Study Design

This study used a cross-sectional survey design. The study was approved by Western Sydney Local Health District Human Research Ethics Committee (Project number 2020/ETH03085).

### Setting

Participants were recruited from 21^st^ March to July 9^th^, 2021. During this period, the COVID-19 vaccine rollout had begun across Australia, and daily cases of community transmission in NSW ranged from 0 – 45(covid19data.com.au, 2021). Restrictions across Greater Sydney began on June 23^rd^, including limitations on the number of people allowed to visit a household, maximum number of people in an exercise class, and reduced seating capacity for outdoor events.(NSW Health, 2021b) On the day the survey closed (July 9^th^) the NSW daily case count was 45, and NSW Health announced stay-at-home orders for Greater Sydney(NSW Health, 2021a). The survey was closed at this time despite some recruitment targets not reached so that results could be more readily interpreted.

### Participants

Participants were eligible to take part if they were aged 18 or over and spoke one of the following as their main language at home: Arabic, Assyrian, Croatian, Dari, Dinka, Hindi, Khmer, Chinese, Samoan/Tongan (combined as one language ‘group’), or Spanish. We selected these ten language groups through iterative discussions with multicultural health staff, with the aim of providing broad coverage across different global regions, groups with varying average levels of English language proficiency (based on 2016 Australian census data)(Australian Bureau of Statistics, 2016), varying access to translated materials, and varying degrees of reading skill in their main language spoken at home (Appendix 2). Each of the language groups selected was an important group within the Greater Western Sydney region (Western Sydney Local Health District, South Western Sydney Local Health District, Nepean Blue Mountains Local Health District).

Participants were recruited through bilingual Multicultural Health staff and Health Care Interpreter Service staff. Multicultural Health staff recruited participants through their existing networks, community events and community champions. Health Care Interpreter Service staff recruited participants at the end of a medical appointment. Potential participants were offered two means of taking part: completing the survey themselves online (available in English or translated), or bilingual staff or an interpreter entered responses into the survey platform on the participants’ behalf. To ensure consistency in the phrases used for assisted survey completion, translated versions of the survey were provided to the bilingual staff and interpreters.

### Survey design

Surveys were available in English or translated, and hosted on the web-based survey platform Qualtrics. Demographic survey items included age, gender, education, years living in Australia, main language spoken at home, self-reported English language proficiency, reading proficiency in language spoken at home, access to the internet, access to smartphones, chronic disease, and a single-item health literacy screener(Wallace et al., 2006).

Participants were asked “*If I get signs of COVID-19 in the next 4 weeks (cough, sore throat, fever), the following might stop me from getting tested,”* with response options adapted from our previous survey findings(McCaffery et al., Bonner et al., 2021), modified to suit the Greater Western Sydney context (e.g. including responses that highlight concerns about visa status). Participants who responded “*I will get tested no matter what*” (and listed no barriers) were coded as high intention to get tested. Three additional items assessed intentions for specific testing behaviours, captured using 5-point Likert scales (strongly disagree to strongly agree). These items asked about intentions to get tested if symptomatic, to stay at home while waiting for results, and to stay home if they experience COVID-19 symptoms. Vaccine willingness was assessed by asking “*If a COVID-19 vaccine were recommended for you, would you get itã*” with response option ‘Yes,’ ‘No,’ or ‘Not sure,’ as recommended by the World Health Organization for assessments of vaccine intention(Organization, 2021). Participants were not asked directly about receipt of a COVID-19 vaccine as we had anticipated recruitment would finish prior to vaccine rollout.

Participants were asked to specify their top 3 information sources for finding out about COVID-19 in the previous 4 weeks. Risk perception was captured by asking “how serious a problem do you think COVID-19 is currently, in Australiaã” with responses ranging from 0 (not serious at all) to 10 (very serious), adapted from our previous COVID-19 surveys(McCaffery et al.).

### Analysis plan

Frequencies were weighted (using post-stratification weighting) to reflect each language group’s gender and age group distribution (18-29 years, 30-49 years, 50-69 years, ≥70 years) based on 2016 census data for the Greater Western Sydney population(Australian Bureau of Statistics, 2016). All summary statistics presented in the results section are weighted unless otherwise indicated. A single participant indicated their gender as ‘other’ and was unable to be included in weighted analyses.

Logistic regression models were used to determine factors associated with testing intentions and vaccine willingness, respectively. Table S3 in the Appendix presents an additional multinomial regression model that examines factors associated with the ‘not sure’ and ‘no’ responses to vaccination compared to ‘yes’ responses; for ease of interpretation we have combined these into a binary variable (‘willing’ vs ‘not sure/no’) in the main body of the manuscript. Age group, gender, health literacy, English-language proficiency, years lived in Australia, risk perception, language group, and information sources were included in each model as these have been identified as correlates in relevant research(Edwards et al., 2021, Dodd et al., 2021). The regression also controlled for socioeconomic status of area of residence (based on Index of Relative Socio-economic Advantage and Disadvantage (IRSAD)(Australian Bureau of Statistics, 2018) deciles by postcode), and whether participants completed the survey before or after 23^rd^ June, when new restrictions were announced for all of Greater Sydney due to a COVID-19 outbreak(NSW Health, 2021b). The IRSAD decile was not available for some participants (n=5), for example, because they had entered digits that did not correspond to a current or previously valid Australian postcode. IRSAD decile for these participants was replaced with the median IRSAD decile for speakers of the same language in the sample. Statistical analysis was conducted using Complex Sample procedures in IBM SPSS Statistics 26.

Free-text responses were analysed using content analysis(Weber, 1990) for the following survey item: “*So that we can understand where there are areas of concern or confusion in the community, please tell us what information you would like to know or don’t understand about COVID-19 (including vaccination)*”. OM familiarised herself with the content and developed a list of preliminary content categories. These categories were refined through discussion with the other authors. OM and CAB coded 534 valid responses according to the final coding framework. The level of agreement was tested using 50 responses, which indicated substantial agreement (Cohen’s κ=0.95)(Landis and Koch, 1977). Discrepancies were discussed with JA before coding the remaining responses.

## Results

### Sample description

The mean respondent age was 45.4 years (95% CI: 43.9 to 47.0; range 18–91 years), and 51% of respondents were female (n=363; Table 1). Most participants (88%, n=622) were born in a country other than Australia; 31% reported that they did not speak English well or at all (n=220); 70% had no tertiary qualifications (n=497). Inadequate health literacy was identified for 41% of the sample (n=290). Mean IRSAD decile was 4.73 (95%CI: 4.47 to 5.00).

**Table 1.** Descriptive statistics (categorical variables)*

| **Variable** | **n** | **%** |
| --- | --- | --- |
| **Age group** |  |  |
| 18-29 | 147 | 20.8 |
| 30-49 | 295 | 41.7 |
| 50-69 | 193 | 27.3 |
| >70 | 72 | 10.2 |
| **Gender\*** |  |  |
| Male | 344 | 48.7 |
| Female | 363 | 51.3 |
| **Language** |  |  |
| Assyrian | 133 | 18.8 |
| Croatian | 121 | 17.1 |
| Arabic | 80 | 11.3 |
| Chinese | 76 | 10.7 |
| Khmer | 63 | 8.9 |
| Dinka | 63 | 8.9 |
| Dari | 44 | 6.2 |
| Spanish<sup>†</sup> | 43 | 6.1 |
| Hindi | 42 | 5.9 |
| Samoan/Tongan | 42 | 5.9 |
| **English language proficiency (How well do you speak English?)** |  |  |
| Very well/ well | 487 | 68.9 |
| Not well/not at all | 220 | 31.1 |
| **Literacy in a language other than English (How well do you read in your main language?)** |  |  |
| Very well/ well | 589 | 83.3 |
| Not well/not at all | 118 | 16.7 |
| **Adequate health literacy** | 417 | 59.0 |
| **Highest level of education** |  |  |
| Less than year 12(less than high school) | 115 | 16.3 |
| Year 12 (high school graduate) | 133 | 18.8 |
| Certificate level I to IV / Advanced diploma and diploma level | 249 | 35.2 |
| Bachelor degree level and above | 210 | 29.7 |
| **Has a computer with internet access** | 573 | 81.0 |
| **Has a smartphone** | 686 | 97.0 |
| **Years living in Australia** |  |  |
| 5 years or less | 120 | 17.0 |
| 6 to 10 years | 104 | 14.7 |
| More than 10 years | 398 | 56.3 |
| Born in Australia | 85 | 12.0 |
| **Chronic health conditions<sup>^</sup>** |  |  |
| 0 | 421 | 59.6 |
| 1 | 154 | 21.8 |
| 2 or more | 132 | 18.6 |
| **Index of Relative Socio-economic Advantage and Disadvantage quintile\*\*** |  |  |
| 1 (Most disadvantaged) | 224 | 31.7 |
| 2 | 131 | 18.6 |
| 3 | 125 | 17.7 |
| 4 | 140 | 19.8 |
| 5 (Least disadvantaged) | 87 | 12.3 |
| **Total** | **707** |  |

\*1 respondent indicated 'other/prefer not to say' and is not included in weighted analysis; <sup>†</sup>Spanish language group had substantial gaps in recruitment across age groups <sup>^</sup>Chronic health conditions included respiratory disease, asthma, chronic obstructive pulmonary disease, high blood pressure, cancer, heart disease, stroke, diabetes, depression or anxiety; \*\*IRSAD presented as quintiles to show overall distribution.

### COVID-19 testing intentions

Overall, participants reported high intentions to get tested in the next four weeks if they experienced COVID-19 symptoms (M=4.4 out of 5, 95% CI: 4.3 to 4.5), stay home while waiting for COVID-19 test results (M=4.5 out of 5, 95% CI:4.4 to 4.6), and to stay home if they experienced symptoms (M=4.6 out of 5, 95% CI: 4.5 to 4.7). Responses did not vary substantially for different demographic groups (Appendix Table S1).

Three quarters of participants (n=546, 77.2%) responded that they would ‘get tested no matter what’ if they developed COVID-19 symptoms in the next four weeks (Table 2). This proportion ranged from 58.7% for Croatian speakers (n=71), to 99.4% for Hindi speakers (n=42). For participants with inadequate health literacy, 72.7% indicated that they would ‘get tested no matter what’ (n=211); compared to 80.3% for people with adequate health literacy (n=335).

**Table 2.** COVID-19 testing intentions and vaccine willingness, descriptive characteristics.

| Variable | COVID-19 testing intention |  | COVID-19 vaccination willingness |  |  |  |  |  |
| --- | --- | --- | --- | --- | --- | --- | --- | --- |
|  | No barriers |  | Yes |  | Not sure |  | No |  |
|  | n | % | n | % | n | % | n | % |
| **Age group** |  |  |  |  |  |  |  |  |
| 18-29 | 108 | 73.8 | 70 | 47.7 | 35 | 23.7 | 42 | 28.7 |
| 30-49 | 236 | 79.8 | 169 | 57.1 | 83 | 28.0 | 44 | 14.9 |
| 50-69 | 155 | 80.2 | 105 | 54.4 | 66 | 34.3 | 22 | 11.2 |
| >70 | 47 | 65.0 | 31 | 42.9 | 21 | 29.7 | 20 | 27.4 |
| **Gender\*** |  |  |  |  |  |  |  |  |
| Male | 262 | 76.1 | 200 | 58.4 | 90 | 26.1 | 53 | 15.5 |
| Female | 284 | 78.2 | 174 | 47.9 | 115 | 31.7 | 74 | 20.4 |
| **Language** |  |  |  |  |  |  |  |  |
| Arabic | 65 | 81.0 | 52 | 64.6 | 8 | 9.6 | 21 | 25.7 |
| Assyrian | 106 | 79.8 | 54 | 40.8 | 27 | 20.5 | 51 | 38.7 |
| Chinese | 57 | 75.5 | 36 | 46.8 | 29 | 38.7 | 11 | 14.5 |
| Croatian | 71 | 58.7 | 53 | 43.5 | 53 | 43.6 | 16 | 12.9 |
| Dari | 33 | 74.4 | 15 | 34.4 | 23 | 53.1 | 6 | 12.5 |
| Dinka | 47 | 74.0 | 31 | 48.8 | 18 | 28.3 | 14 | 22.9 |
| Hindi | 42 | 99.4 | 34 | 81.9 | 8 | 18.1 | 0 | 0.0 |
| Khmer | 56 | 88.9 | 62 | 98.4 | 1 | 1.6 | 0 | 0.0 |
| Spanish<sup>†</sup> | 40 | 93.3 | 26 | 59.9 | 14 | 32.6 | 3 | 7.5 |
| Samoan/Tongan | 29 | 69.0 | 12 | 29.4 | 24 | 57.6 | 5 | 12.9 |
| **English language proficiency** |  |  |  |  |  |  |  |  |
| Very well/ well | 384 | 78.9 | 265 | 54.3 | 129 | 26.4 | 94 | 19.3 |
| Not well/not at all | 161 | 73.4 | 110 | 49.9 | 77 | 34.8 | 34 | 15.3 |
| **Literacy in a language other than English** |  |  |  |  |  |  |  |  |
| Very well/ well | 452 | 76.7 | 305 | 51.7 | 189 | 32.0 | 96 | 16.3 |
| Not well/not at all | 94 | 79.5 | 70 | 59.3 | 16 | 14.0 | 32 | 26.8 |
| **Health literacy** |  |  |  |  |  |  |  |  |
| Adequate | 335 | 80.3 | 235 | 56.4 | 112 | 26.8 | 70 | 16.8 |
| Inadequate | 211 | 72.7 | 139 | 48.0 | 93 | 32.2 | 57 | 19.8 |
| **Years living in Australia** |  |  |  |  |  |  |  |  |
| 5 years or less | 91 | 76.0 | 75 | 62.9 | 24 | 19.8 | 21 | 17.3 |
| 6 to 10 years | 78 | 75.4 | 63 | 61.0 | 32 | 30.7 | 9 | 8.3 |
| More than 10 years | 305 | 76.7 | 198 | 49.8 | 130 | 32.7 | 70 | 17.5 |
| Born in Australia | 71 | 83.4 | 37 | 44.0 | 19 | 22.6 | 28 | 33.4 |
| **Chronic health conditions<sup>^</sup>** |  |  |  |  |  |  |  |  |
| 0 | 338 | 80.3 | 239 | 56.9 | 78 | 18.6 | 103 | 24.5 |
| 1 | 107 | 69.3 | 80 | 51.5 | 19 | 12.4 | 56 | 36.0 |
| 2 or more | 101 | 76.5 | 56 | 42.2 | 30 | 22.7 | 46 | 35.1 |
| **Index of Relative Socio-economic Advantage and Disadvantage (quintile)\*\*** |  |  |  |  |  |  |  |  |
| 1 (Most disadvantaged) | 183 | 81.6 | 117 | 52.1 | 54 | 24.1 | 53 | 23.8 |
| 2 | 101 | 77.2 | 83 | 63.0 | 34 | 25.6 | 15 | 11.4 |
| 3 | 78 | 62.1 | 57 | 45.2 | 43 | 34.2 | 26 | 20.6 |
| 4 | 107 | 76.2 | 71 | 51.1 | 44 | 31.2 | 25 | 17.7 |
| 5 (Least disadvantaged) | 77 | 88.9 | 47 | 54.4 | 31 | 35.8 | 9 | 9.8 |
| **Total** | **546** | **77.2** | **375** | **53.0** | **205** | **29.0** | **127** | **18.0** |

\* 1 respondent indicated ‘other/prefer not to say’ and is not included in weighted analysis; <sup>†</sup>Spanish language group had substantial gaps in recruitment across age groups; <sup>^</sup>Chronic health conditions included respiratory disease, asthma, chronic obstructive pulmonary disease, high blood pressure, cancer, heart disease, stroke, diabetes, depression or anxiety; \*\*IRSAD presented as quintiles to show overall distribution.

The most frequently reported information source for participants who said they would get tested for COVID-19 if they had symptoms ‘no matter what,’ and for participants who listed barriers to testing was official Australian sources or public broadcasters, followed by Australian commercial sources, and social media (Appendix Table S2). Two fifths of participants who listed barriers to testing reporting using overseas information sources (41.4% vs no barriers: 25.5%) community sources (38.0% vs no barriers: 25.3%), or family or friends living in Australia (40.5% vs no barriers 34.2%) as a main way of finding out about COVID-19.

Intention to get tested for COVID-19 symptoms ‘no matter what’ was significantly associated with age (p=0.03), with participants aged 50-69 years less likely to get tested than participants aged less than 30 years (OR=0.44, 95% CI: 0.20 to 0.99, p<0.001) (Table 3). Intention to get tested also varied significantly across language groups (p<0.001). No information sources were significantly associated with testing intention once adjusted for other covariates.

**Table 3.**
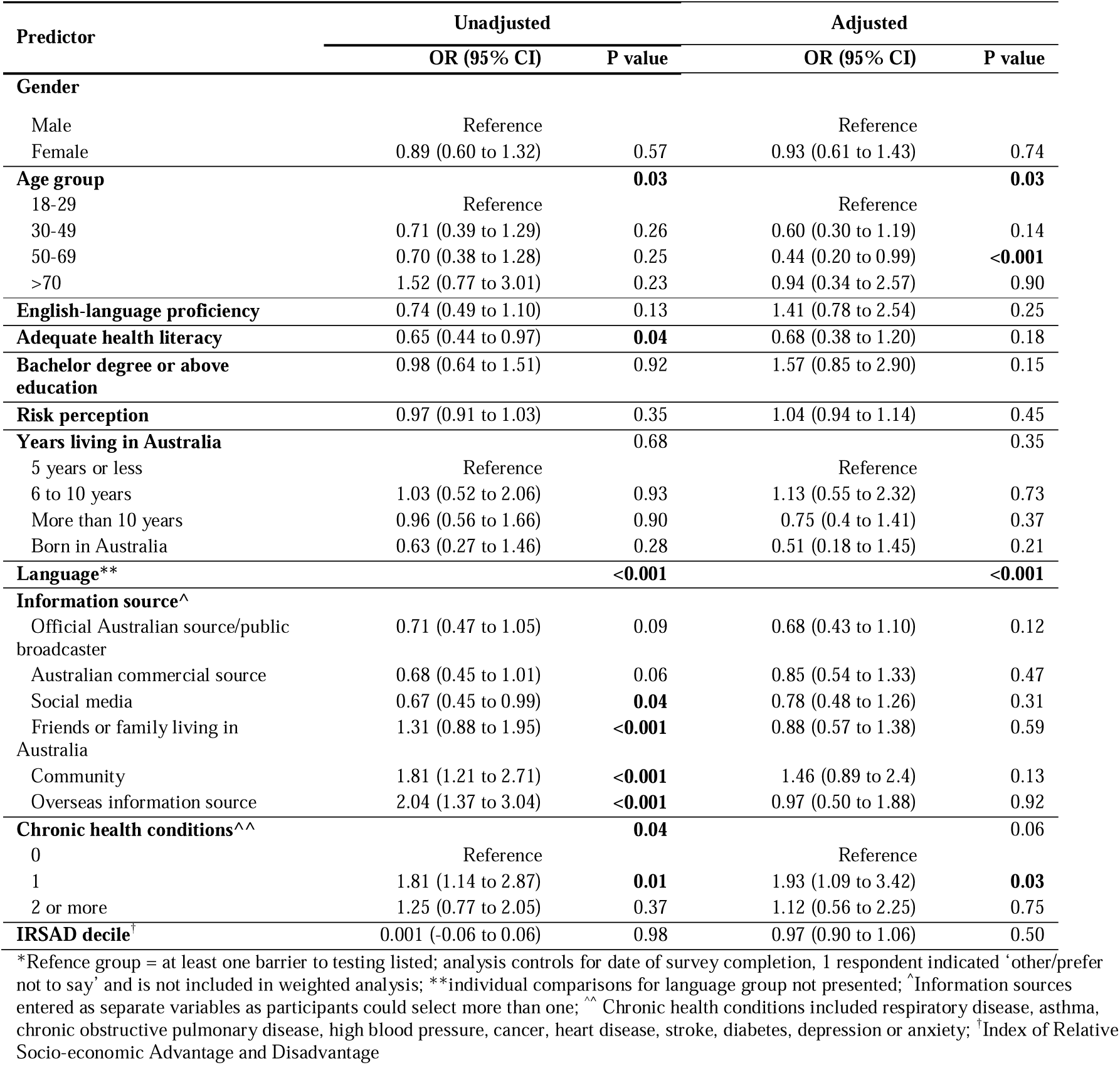
Logistic regression model predicting COVID-19 testing intentions (‘I will get tested no matter what’)*

| Predictor | Unadjusted |  | Adjusted |  |
| --- | --- | --- | --- | --- |
|  | OR (95% CI) | P value | OR (95% CI) | P value |
| **Gender** |  |  |  |  |
| Male | Reference |  | Reference |  |
| Female | 0.89 (0.60 to 1.32) | 0.57 | 0.93 (0.61 to 1.43) | 0.74 |
| **Age group** |  | **0.03** |  | **0.03** |
| 18-29 | Reference |  | Reference |  |
| 30-49 | 0.71 (0.39 to 1.29) | 0.26 | 0.60 (0.30 to 1.19) | 0.14 |
| 50-69 | 0.70 (0.38 to 1.28) | 0.25 | 0.44 (0.20 to 0.99) | **<0.001** |
| >70 | 1.52 (0.77 to 3.01) | 0.23 | 0.94 (0.34 to 2.57) | 0.90 |
| **English-language proficiency** | 0.74 (0.49 to 1.10) | 0.13 | 1.41 (0.78 to 2.54) | 0.25 |
| **Adequate health literacy** | 0.65 (0.44 to 0.97) | **0.04** | 0.68 (0.38 to 1.20) | 0.18 |
| **Bachelor degree or above education** | 0.98 (0.64 to 1.51) | 0.92 | 1.57 (0.85 to 2.90) | 0.15 |
| **Risk perception** | 0.97 (0.91 to 1.03) | 0.35 | 1.04 (0.94 to 1.14) | 0.45 |
| **Years living in Australia** |  | 0.68 |  | 0.35 |
| 5 years or less | Reference |  | Reference |  |
| 6 to 10 years | 1.03 (0.52 to 2.06) | 0.93 | 1.13 (0.55 to 2.32) | 0.73 |
| More than 10 years | 0.96 (0.56 to 1.66) | 0.90 | 0.75 (0.4 to 1.41) | 0.37 |
| Born in Australia | 0.63 (0.27 to 1.46) | 0.28 | 0.51 (0.18 to 1.45) | 0.21 |
| **Language\*\*** |  | **<0.001** |  | **<0.001** |
| **Information source^** |  |  |  |  |
| Official Australian source/public broadcaster | 0.71 (0.47 to 1.05) | 0.09 | 0.68 (0.43 to 1.10) | 0.12 |
| Australian commercial source | 0.68 (0.45 to 1.01) | 0.06 | 0.85 (0.54 to 1.33) | 0.47 |
| Social media | 0.67 (0.45 to 0.99) | **0.04** | 0.78 (0.48 to 1.26) | 0.31 |
| Friends or family living in Australia | 1.31 (0.88 to 1.95) | **<0.001** | 0.88 (0.57 to 1.38) | 0.59 |
| Community | 1.81 (1.21 to 2.71) | **<0.001** | 1.46 (0.89 to 2.4) | 0.13 |
| Overseas information source | 2.04 (1.37 to 3.04) | **<0.001** | 0.97 (0.50 to 1.88) | 0.92 |
| **Chronic health conditions^^** |  | **0.04** |  | 0.06 |
| 0 | Reference |  | Reference |  |
| 1 | 1.81 (1.14 to 2.87) | **0.01** | 1.93 (1.09 to 3.42) | **0.03** |
| 2 or more | 1.25 (0.77 to 2.05) | 0.37 | 1.12 (0.56 to 2.25) | 0.75 |
| **IRSAD decile†** | 0.001 (-0.06 to 0.06) | 0.98 | 0.97 (0.90 to 1.06) | 0.50 |

\*Refence group = at least one barrier to testing listed; analysis controls for date of survey completion, 1 respondent indicated 'other/prefer not to say' and is not included in weighted analysis; \*\*individual comparisons for language group not presented; ^Information sources entered as separate variables as participants could select more than one; ^^ Chronic health conditions included respiratory disease, asthma, chronic obstructive pulmonary disease, high blood pressure, cancer, heart disease, stroke, diabetes, depression or anxiety; †Index of Relative Socio-economic Advantage and Disadvantage

Of the 161 participants who identified at least one barrier to getting tested for COVID-19 in the next four weeks, the most common barrier to testing was concern about infection at the testing clinic (26.1%, n=42) (Appendix Tables S3-S4). This was followed by concerns that testing is painful (25.3%, n=41), and the belief that if they had already received a negative test, they did not need or want another one (13.9%, n=22).

### COVID-19 vaccine willingness

In total, 53.0% of participants (n=375) said that they would get a COVID-19 vaccine if it was recommended to them, 29.0% (n=205) indicated they were not sure, and 18.0% (n=127) responded that they would not get the vaccine (Table 2). Vaccine willingness (responding ‘yes’) was 58.4% (n=200) for male participants, compared to 47.9% (n=174) for female participants. This proportion was 56.5% for people with adequate health literacy (n=235) compared to 47.8% for people with inadequate health literacy (n=139). Across language groups, vaccine acceptance ranged from 29.5% (Samoan/Tongan, n=12) to 98.4% (Khmer, n=62) (Table 2). Responding ‘not sure’ was highest for Samoan/Tongan (57.6%, n=24) and Dari speakers (53.2%, n=23), whereas responding ‘no’ was highest for Assyrian (38.7%, n=51) and Arabic speakers (25.8%, n=21).

For participants who were accepting of the vaccine, the main information sources were official Australian sources or public broadcasters (62.7%, n=235), followed by Australian commercial sources (67.1%, n=251), and social media (58.2%, n=218) (Appendix Table S2). Participants who responded ‘no’ or ‘not sure’ reported the same top three information sources.

We observed significant differences in vaccination willingness across language groups (p<0.001), controlling for all other covariates (Table 4). Female participants were also more likely to be hesitant about COVID-19 vaccination (OR= 1.63, 95% CI: 1.10 to 2.43, p=0.02), as were participants who did not use Australian commercial information sources as a main way of finding out about COVID-19 (OR= 0.57, 95%CI: 0.37 to 0.86, p=0.01). No other differences were observed across sociodemographic variables.

**Table 4.** Logistic regression model predicting COVID-19 vaccination willingness*.

| Predictor | Not sure/No (vs yes willing) |  |  |  |
| --- | --- | --- | --- | --- |
|  | Unadjusted |  | Adjusted |  |
|  | OR (95% CI) | P value | OR (95% CI) | P value |
| **Gender** |  |  |  |  |
| Male | Reference |  | Reference |  |
| Female | 1.52 (1.06 to 2.18) | **0.02** | 1.63 (1.10 to 2.43) | **0.02** |
| **Age group** |  | 0.16 |  | 0.52 |
| 18-29 | Reference |  | Reference |  |
| 30-49 | 0.68 (0.39 to 1.21) | 0.19 | 0.70 (0.37 to 1.32) | 0.27 |
| 50-69 | 0.76 (0.43 to 1.36) | 0.36 | 0.58 (0.28 to 1.19) | 0.14 |
| >70 | 1.21 (0.61 to 2.39) | 0.58 | 0.62 (0.25 to 1.55) | 0.31 |
| **English-language proficiency** | 0.84 (0.59 to 1.20) | 0.33 | 0.93 (0.53 to 1.63) | 0.80 |
| **Adequate health literacy** | 0.71 (0.50 to 1.03) | 0.07 | 0.78 (0.47 to 1.28) | 0.33 |
| **Bachelor degree or above education** | 0.89 (0.60 to 1.32) | 0.57 | 1.09 (0.64 to 1.86) | 0.75 |
| **Risk perception** | 0.95 (0.90 to 1.01) | 0.11 | 1.00 (0.92 to 1.08) | 0.91 |
| **Years living in Australia** |  | 0.06 |  | 0.18 |
| 5 years or less | Reference |  | Reference |  |
| 6 to 10 years | 1.08 (0.57 to 2.06) | 0.81 | 1.43 (0.70 to 2.95) | 0.33 |
| More than 10 years | 1.71 (1.03 to 2.82) | 0.04 | 1.85 (0.97 to 3.52) | 0.06 |
| Born in Australia | 2.16 (0.95 to 4.88) | 0.07 | 2.34 (0.97 to 5.65) | 0.06 |
| **Language\*\*** |  | **<0.001** |  | **<0.001** |
| **Information source^^** |  |  |  |  |
| Official Australian source/public broadcaster | 0.78 (0.54 to 1.12) | 0.18 | 1.15 (0.72 to 1.81) | 0.56 |
| Australian commercial source | 0.50 (0.34 to 0.71) | **<0.001** | 0.57 (0.37 to 0.86) | **0.01** |
| Social media | 0.84 (0.59 to 1.21) | 0.35 | 0.89 (0.59 to 1.35) | 0.59 |
| Friends or family living in Australia | 1.43 (0.98 to 2.10) | 0.06 | 1.15 (0.75 to 1.76) | 0.52 |
| Community | 1.10 (0.76 to 1.59) | 0.63 | 0.95 (0.58 to 1.55) | 0.83 |
| Overseas information source | 1.09 (0.76 to 1.57) | 0.63 | 0.60 (0.34 to 1.06) | 0.08 |
| **Chronic health conditions^** |  | **0.04** |  | 0.19 |
| 0 | Reference |  | Reference |  |
| 1 | 1.24 (0.81 to 1.89) | 0.32 | 1.33 (0.78 to 2.29) | 0.30 |
| 2 or more | 1.81 (1.15 to 2.84) | **0.01** | 1.68 (0.95 to 2.97) | 0.08 |
| **IRSAD decile†** | 1.01 (0.95 to 1.08) | 0.76 | 1.02 (0.93 to 1.11) | 0.66 |

\*Refence group = 'yes' (i.e. willing to get a vaccine); analysis controls for IRSAD and date of survey completion, 1 respondent indicated 'other/prefer not to say' and is not included in weighted analysis; \*\*individual comparisons for language group not presented; ^Information sources entered as separate variables as participants could select more than one; ^^ Chronic health conditions included respiratory disease, asthma, chronic obstructive pulmonary disease, high blood pressure, cancer, heart disease, stroke, diabetes, depression or anxiety; † p value for test of model effect includes multinomial regression (both 'not sure' and 'no' responses to vaccine intention); † Index of Relative Socio-economic Advantage and Disadvantage

Of the 335 participants who were not accepting of the vaccine (no/not sure), almost one third (30.7%, n=102) indicated concern about vaccine side effects as their main reason (Appendix Table S6). A further 24.2% (n=80) listed safety as a concern, and 11.5% (n=38) needed more information before deciding. This ranking was similar across language groups (Appendix Table S7).

Three quarters of participants (76.3%, n=534) provided free-text responses identifying areas of concern or confusion about COVID-19. We generated 11 topics to categorise responses (Table 5). More than half (53.0%, n=286) voiced concerns about the COVID-19 vaccines, with 23.9% (n=129) specifically concerned about vaccine safety and side effects. Almost one fifth voiced concerns about how COVID-19 information was communicated (17.4%, n=94), discussing the need for translated information (8.4%), clearer and more accurate information (8.2%), or information available in different formats (3.2%).

**Table 5.** Content analysis of areas of concern or confusion about COVID-19 in the community*.

| Area of concern or confusion | Example quote | N | % |
| --- | --- | --- | --- |
| **Information and communication** | *"Clearer to the point information not an overload;* | **94** | **17.4** |
| Need for translated information | *currently there is too much information available that is* | 45 | **8.4** |
| Need for clear, accurate, and high quality information | *badly translated and not targeting my community* | 44 | **8.2** |
| Need for information available in different formats | *appropriately it is written for highly educated and it is too wordy"* | 17 | **3.2** |
| **Vaccines** | *"Doubts about vaccines. No clarity from government on* | **286** | **53.0** |
| Vaccine safety and side effects | *reactions and side effects. Still unclear about my eligibility* | 129 | **23.9** |
| General vaccine questions | *for vaccinations. Do we have to take every year like flu* | 86 | **16.0** |
| Vaccine rollout and practical barriers | *injections- no clear information."* | 58 | **10.7** |
| Vaccine efficacy |  | 30 | **5.6** |
| Concerns about AstraZeneca | *"Worried about blood clots post vaccine"* | 20 | **3.8** |
| **Other** | *"The type of Covid-19 and the time it is going to end"* | 116 | **21.4** |
| General covid/pandemic concerns and questions |  | 72 | **13.4** |
| Need more information about travel bans and border restrictions | *"For me, being away from my children is very difficult. I want to know for how long should we remain away from our families"* | 30 | **5.6** |
| Other |  | 15 | **2.7** |
| **No concerns or areas of confusion** | *"no question as my doctor will inform me on Covid"* | **95** | **17.7** |

\*1 respondent indicated 'other/prefer not to say' and is not included in weighted analysis; responses could be coded to more than one area of concern or confusion.

## Discussion

### Key findings

This study provides insight into the perspectives of culturally and linguistically diverse Australians during the start of the COVID-19 vaccine rollout, a period characterised by low levels of COVID-19 community transmission (March 21 to July 9, 2021). Three quarters of participants (77%) reported that they would get tested for COVID-19 if they experienced symptoms, ‘no matter what,’ though this proportion varied significantly across language groups. The main barriers to testing were concerns about getting infected at the testing centre and about the test being painful. Just over half the sample (53%) were willing to get a COVID-19 vaccine if recommended to them, one fifth responded they were unwilling, and the remainder unsure, though again, these proportions varied significantly by language spoken at home. Participants were more likely to say ‘no’ or ‘not sure’ to a COVID-19 vaccine if they were female or did not use Australian commercial information sources as a main way of finding out about COVID-19. Key barriers to vaccination were concerns about side effects and safety of COVID-19 vaccines, and a need for more information before making a decision.

We observed higher testing intentions in this study compared to national Australian estimates during a similar time period (77% vs 45%-51%)(Australian Bureau of Statistics, 2021a, Australian Bureau of Statistics, 2021b), although different survey items were used. This study identified different top barriers to testing as in our previous Australia-wide survey in November 2020; though both observed concerns about the test being painful within the top 5 barriers(Bonner et al., 2021).

Conversely, we observed lower COVID-19 vaccine willingness in this study compared to national Australian estimates during similar time periods (53% vs 68%-73%)(Australian Bureau of Statistics, 2021a, Australian Bureau of Statistics, 2021b), though this rate was similar to a study of 199 people from NSW, Australia, who spoke a language other than English at home (58%)(NSW Council of Social Services, 2021). Key barriers to vaccination were similar in this study and our November Australia-wide survey (i.e. concerns about safety and needing more information)(Dodd et al., 2021), though distrust of the government did not feature highly in the current study; only 3% of participants who reported barriers to vaccination said this was because they did not trust the government. This is consistent with findings from a survey of 656 Australian refugees and asylum seekers, in which trust in authorities was not identified as a key COVID-19 concern(Liddell et al., 2021).

In this study the main factor associated with testing and vaccination intentions was the language spoken at home, even when controlling for education, socioeconomic status, and English-language proficiency. Given existing inequities for people from culturally and linguistically diverse communities, and the lower vaccination intentions observed in this study, public health should prioritise these groups to support testing and vaccination(Dodds and Fakoya, 2020, Al-Oraibi et al., 2021). This approach must work collaboratively with people in these communities to develop tailored, targeted approaches to COVID-19 vaccine communication and rollout(Wild et al., 2021, Leask et al., 2021, Chauhan et al., 2021, Raz et al., 2021). In fact, the Australian federal government itself laid out policy in November 2020(Australian Government, 2021a), emphasising the need for translated and simple English communication about the vaccines and their rollout, providing ample opportunity for people in these communities to ask questions, working with community leaders and representatives, and embedding interpreter workforce into clinical services. However, at the time of study recruitment this policy had not been adequately implemented in Australia. There is evidence that this inclusive approach can also support COVID-19 testing behaviours, as evidenced by community-based strategies implemented in culturally and linguistically diverse neighbourhoods in Seattle in the US(Kim et al., 2020). This study also supports recent commentary reminding researchers that when collective terms for culturally and linguistically diverse populations are not accompanied by disaggregated outputs, they may inadvertently imply homogeneity where it does not exist(Routen et al., 2021).

### Strengths and limitations

The main strength of this study is that we addressed deficiencies in previous research by using more inclusive recruitment and data collection methods to increase opportunity for participation. This included providing translated versions of the survey, using interpreters, and multiple recruitment methods (including through social media, community events, and through community networks). We also included several variables related to culture and language (e.g. English language proficiency, literacy in own language, and years living in Australia), and focused on 10 specific language groups, in an attempt to provide a more nuanced description of the sample that captures some of the complexity and diversity within these communities. Whilst this inevitably means that not all cultural and language groups are represented in the survey, we hope that that by focusing on 10 groups with different access to translated materials, English language skills and proficiency in language spoken at home, that the findings have practical value and can inform decision-making to develop tailored supports and resources that serve these communities. This study also did not ask participants if they had already received the vaccine. State-wide estimates suggest fewer than 25 per 100 NSW population had received a single dose of the vaccine at the time recruitment closed(covid19data.com.au, 2021). Lastly, we were unable to incorporate specific items about the Astrazeneca vaccine as recruitment began before the news about its side effects hit mainstream media outlets.

## Conclusions

This study of people from culturally and linguistically diverse communities in Greater Western Sydney shows that community members have genuine concerns about COVID-19 testing and vaccination, particularly with regards to safety and wanting more information. Different language groups have unique and specific needs that must be met through local targeted communication strategies. Public health bodies must work collaboratively with communities to provide tailored approaches that support people in these communities to take up COVID-19 testing and vaccination, or risk exacerbating health inequalities further.

## Data Availability

Data produced in the present study can be made available upon reasonable request to the authors

## Acknowledgements

We would like to acknowledge the efforts of all community health workers, local health district staff, community champions, and community leaders who supported and contributed to this project. We would also like to thank all participants for their involvement.

## Supplementary materials

Appendix 1: Supplementary tables

Appendix 2: Language selection details

