## Appendix Table for "COVID-19 testing and vaccine willingness: Cross-sectional survey in a culturally diverse community in Sydney, Australia"

### Appendix – Supplementary tables

All N in tables below are weighted for age group and gender distribution according to 2016 Census data. Regression models use the complex samples procedure in SPSS (i.e. also weighted).

**Table S1. Intentions for behaviours relevant to COVID-19 testing***

|  | **Get tested if experiencing COVID-19 symptoms** | | **Stay home while waiting for COVID-19 test results** | | **Stay home if experiencing COVID-19 symptoms** | |
| --- | --- | --- | --- | --- | --- | --- |
| **Variable** | **M** | **SE** | **M** | **SE** | **M** | **SE** |
| **Age group** |  |  |  |  |  |  |
| 18-29 | 4.4 | 0.13 | 4.5 | 0.13 | 4.7 | 0.07 |
| 30-49 | 4.4 | 0.09 | 4.5 | 0.08 | 4.6 | 0.08 |
| 50-69 | 4.4 | 0.10 | 4.5 | 0.11 | 4.6 | 0.11 |
| >70 | 4.4 | 0.09 | 4.5 | 0.08 | 4.5 | 0.08 |
| **Gender*** |  |  |  |  |  |  |
| Male | 4.4 | 0.08 | 4.5 | 0.08 | 4.6 | 0.07 |
| Female | 4.4 | 0.07 | 4.5 | 0.07 | 4.6 | 0.06 |
| **Language** |  |  |  |  |  |  |
| Arabic | 4.6 | 0.14 | 4.7 | 0.13 | 4.8 | 0.11 |
| Assyrian | 4.3 | 0.11 | 4.5 | 0.09 | 4.7 | 0.07 |
| Chinese | 4.8 | 0.05 | 4.7 | 0.20 | 4.9 | 0.05 |
| Croatian | 4.2 | 0.07 | 4.3 | 0.07 | 4.3 | 0.07 |
| Dari | 4.6 | 0.23 | 4.9 | 0.05 | 4.9 | 0.03 |
| Dinka | 4.6 | 0.13 | 4.8 | 0.11 | 4.7 | 0.13 |
| Hindi | 4.3 | 0.32 | 4.0 | 0.33 | 4.1 | 0.31 |
| Khmer | 4.9 | 0.04 | 4.9 | 0.04 | 4.9 | 0.04 |
| Spanish** | 4.3 | 0.19 | 4.3 | 0.16 | 4.8 | 0.10 |
| Samoan/Tongan | 3.4 | 0.37 | 3.5 | 0.40 | 3.8 | 0.45 |
| **English language proficiency** |  |  |  |  |  |  |
| Very well/ well | 4.4 | 0.07 | 4.5 | 0.07 | 4.6 | 0.07 |
| Not well/not at all | 4.5 | 0.07 | 4.6 | 0.05 | 4.6 | 0.05 |
| **Literacy in a language other than English** |  |  |  |  |  |  |
| Very well/ well | 4.4 | 0.06 | 4.5 | 0.06 | 4.6 | 0.06 |
| Not well/not at all | 4.5 | 0.10 | 4.5 | 0.10 | 4.7 | 0.08 |
| **Health literacy** |  |  |  |  |  |  |
| Adequate | 4.5 | 0.07 | 4.5 | 0.07 | 4.7 | 0.06 |
| Inadequate | 4.4 | 0.09 | 4.4 | 0.09 | 4.5 | 0.08 |
| **Years living in Australia** |  |  |  |  |  |  |
| 5 years or less | 4.7 | 0.10 | 4.7 | 0.07 | 4.8 | 0.07 |
| 6 to 10 years | 4.5 | 0.16 | 4.6 | 0.16 | 4.7 | 0.15 |
| More than 10 years | 4.4 | 0.07 | 4.4 | 0.07 | 4.5 | 0.07 |
| Born in Australia | 4.2 | 0.17 | 4.4 | 0.18 | 4.7 | 0.09 |
| **Total** | 4.4 | 0.05 | 4.5 | 0.05 | 4.6 | 0.05 |

*Each item rated on a 5-point likert scale, from 1 (strongly disagree) to 5 (strongly agree); 1 respondent indicated ‘other/prefer not to say’ and is not included in weighted analysis; **Spanish language group had substantial gaps in recruitment across age groups

**Table S2. Main COVID-19 information sources, by testing intention and vaccination willingness***

| **Main COVID-19 information sources** | **COVID-19 testing intention** | | | | **COVID-19 vaccination willingness** | | | | | |
| --- | --- | --- | --- | --- | --- | --- | --- | --- | --- | --- |
|  | **No barriers** | | **≥1 barriers** | | **Yes** | | **Not sure** | | **No** | |
|  | **N** | **%** | **N** | **%** | **N** | **%** | **N** | **%** | **N** | **%** |
| Official Australian source / public broadcaster | 337 | 61.8 | 86 | 53.3 | 235 | 62.7 | 117 | 57.0 | 71 | 56.0 |
| Australian Commercial | 336 | 61.6 | 84 | 52.0 | 252 | 67.3 | 109 | 52.9 | 59 | 46.5 |
| Social media | 322 | 58.9 | 79 | 48.9 | 220 | 58.6 | 122 | 59.3 | 59 | 46.5 |
| Friends or family living in Australia | 187 | 34.2 | 65 | 40.5 | 119 | 31.8 | 87 | 42.3 | 46 | 36.3 |
| Community | 138 | 25.3 | 61 | 38.0 | 102 | 27.3 | 70 | 34.2 | 27 | 21.2 |
| Overseas sources | 139 | 25.5 | 66 | 41.1 | 106 | 28.2 | 77 | 37.3 | 23 | 18.3 |

* 1 respondent indicated ‘other/prefer not to say’ and is not included in weighted analysis; Participants could select more than one main information source;

**Table S3. Barriers to COVID-19 testing***

| **Barrier** | **n** | **%** |
| --- | --- | --- |
| I'm worried I will get infected with COVID-19 at the testing clinic | 42 | 26.1 |
| Testing is painful | 41 | 25.3 |
| I've already had a test and it came back negative, so I don't need or want to get another one | 22 | 13.9 |
| I'll forget to get tested | 21 | 12.8 |
| I don't know how, when, and where to get tested | 18 | 11.1 |
| I'm worried that the results will be on my health record | 17 | 10.8 |
| I'm worried about what will happen to my visa if I test positive | 12 | 7.6 |
| I'm worried about what others think of me | 12 | 7.4 |
| Other (E.g. will only get tested for severe symptoms) | 9 | 5.7 |
| It's too difficult or expensive to get tested | 6 | 3.8 |

*1 respondent indicated ‘other/prefer not to say’ and is not included in weighted analysis; more than one barrier could be selected.

**Table S4. Barriers to COVID-19 testing, by language group***

| **Barriers** | **Arabic** | | **Assyrian** | | **Croatian** | | **Dari** | | **Dinka** | | **Hindi** | | **Khmer** | | **Chinese** | | **Samoan/ Tongan** | | **Spanish^^^** | |
| --- | --- | --- | --- | --- | --- | --- | --- | --- | --- | --- | --- | --- | --- | --- | --- | --- | --- | --- | --- | --- |
|  | **n** | **%** | **n** | **%** | **n** | **%** | **n** | **%** | **n** | **%** | **n** | **%** | **n** | **%** | **n** | **%** | **n** | **%** | **n** | **%** |
| I'm worried I will get infected with COVID-19 at the testing clinic | 1 | 6.9 | 11 | 42.8 | 2 | 4.8 | 2 | 13.5 | 6 | 35.2 | 0 | 0.0 | 4 | 62.2 | 12 | 64.2 | 3 | 20.7 | 1 | 31.5 |
| Testing is painful | 5 | 29.8 | 9 | 33.0 | 4 | 7.7 | 1 | 4.9 | 10 | 59.2 | 0 | 100.0 | 1 | 19.4 | 1 | 7.9 | 9 | 69.6 | 1 | 40.8 |
| I've already had a test and it came back negative, so I don't need or want to get another one | 2 | 15.5 | 2 | 6.0 | 7 | 14.3 | 0 | 0.0 | 4 | 27.0 | 0 | 0.0 | 0 | 0.0 | 4 | 18.9 | 3 | 19.4 | 1 | 27.8 |
| I'll forget to get tested | 1 | 4.0 | 0 | 0.0 | 16 | 31.1 | 0 | 0.0 | 4 | 27.2 | 0 | 0.0 | 0 | 0.0 | 0 | 0.0 | 0 | 0.0 | 0 | 0.0 |
| I don't know how, when, and where to get tested | 3 | 18.1 | 3 | 11.8 | 1 | 1.9 | 4 | 39.5 | 1 | 5.9 | 0 | 0.0 | 0 | 6.2 | 4 | 21.3 | 1 | 9.4 | 0 | 0.0 |
| I'm worried about what will happen to my visa if I test positive | 0 | 2.9 | 0 | 0.0 | 12 | 23.9 | 3 | 23.4 | 0 | 0.5 | 0 | 0.0 | 2 | 24.5 | 1 | 3.3 | 0 | 0.0 | 0 | 0.0 |
| I'm worried that the results will be on my health record | 2 | 10.2 | 1 | 2.9 | 7 | 13.7 | 1 | 8.9 | 1 | 5.9 | 0 | 0.0 | 0 | 0.0 | 0 | 0.0 | 2 | 12.8 | 0 | 0.0 |
| I'm worried about what others think of me | 0 | 0.0 | 3 | 12.1 | 4 | 8.4 | 1 | 8.7 | 3 | 16.1 | 0 | 0.0 | 0 | 0.0 | 1 | 4.5 | 0 | 0.0 | 0 | 0.0 |
| Other | 4 | 25.6 | 3 | 12.5 | 0 | 0.0 | 0 | 0.0 | 1 | 6.8 | 0 | 0.0 | 0 | 0.0 | 1 | 4.5 | 0 | 0.0 | 0 | 0.0 |
| It's too difficult or expensive to get tested | 0 | 0.0 | 2 | 9.2 | 1 | 2.4 | 2 | 14.4 | 0 | 0.0 | 0 | 100.0 | 0 | 0.0 | 1 | 3.4 | 0 | 0.0 | 0 | 0.0 |

*1 respondent indicated ‘other/prefer not to say’ and is not included in weighted analysis; more than one barrier could be selected. ^^^Spanish language group had substantial gaps in recruitment across age groups

**Table S5. Logistic regression model predicting COVID-19 vaccination willingness***

|  | **Not sure (vs yes willing)** | | | | **No (vs yes willing)** | | | | **Test of model effect** | |
| --- | --- | --- | --- | --- | --- | --- | --- | --- | --- | --- |
| **Predictor** | **Unadjusted** | | **Adjusted** | | **Unadjusted** | | **Adjusted** | | **Unadjusted** | **Adjusted** |
|  | **OR (95% CI)** | **P value** | **OR (95% CI)** | **P value** | **OR (95% CI)** | **P value** | **OR (95% CI)** | **P value** | **P value**^†^ | **P value**^†^ |
| **Gender** |  |  |  |  |  |  |  |  | 0.07 | 0.07 |
| Male | Reference |  | Reference |  | Reference |  | Reference |  |  |  |
| Female | 1.48 (0.99 to 2.21) | 0.05 | 1.52 (0.98 to 2.37) | 0.06 | 1.60 (0.94 to 2.7) | 0.08 | 1.73 (0.99 to 3.00) | 0.05 |  |  |
| **Age group** |  |  |  |  |  |  |  |  | **0.01** | 0.07 |
| 18-29 | Reference |  | Reference |  | Reference |  | Reference |  |  |  |
| 30-49 | 0.99 (0.51 to 1.91) | 0.97 | 0.93 (0.45 to 1.92) | 0.84 | 0.43 (0.20 to 0.92) | **0.03** | 0.50 (0.21 to 1.19) | 0.12 |  |  |
| 50-69 | 1.27 (0.66 to 2.46) | 0.48 | 0.90 (0.39 to 2.05) | 0.80 | 0.34 (0.16 to 0.74) | **0.01** | 0.29 (0.11 to 0.76) | 0.01 |  |  |
| >70 | 1.39 (0.63 to 3.07) | 0.41 | 0.65 (0.22 to 1.92) | 0.43 | 1.06 (0.46 to 2.48) | 0.89 | 0.73 (0.21 to 2.52) | 0.61 |  |  |
| **English-language proficiency** | 0.70 (0.47 to 1.04) | 0.08 | 0.79 (0.42 to 1.5) | 0.47 | 1.16 (0.69 to 1.96) | 0.58 | 1.38 (0.63 to 3.01) | 0.42 | 0.11 | 0.43 |
| **Adequate health literacy** | 0.71 (0.48 to 1.05) | 0.09 | 0.83 (0.48 to 1.45) | 0.51 | 0.72 (0.42 to 1.24) | 0.24 | 0.73 (0.37 to 1.43) | 0.36 | 0.18 | 0.61 |
| **Bachelor degree or above education** | 0.88 (0.56 to 1.38) | 0.57 | 1.24 (0.67 to 2.27) | 0.49 | 0.92 (0.53 to 1.60) | 0.76 | 0.82 (0.4 to 1.69) | 0.59 | 0.84 | 0.57 |
| **Risk perception** | 0.98 (0.92 to 1.05) | 0.60 | 1 (0.91 to 1.09) | 0.93 | 0.90 (0.84 to 0.98) | **0.01** | 1 (0.9 to 1.11) | 0.95 | **0.04** | 0.99 |
| **Years living in Australia** |  |  |  |  |  |  |  |  | **0.03** | 0.25 |
| 5 years or less |  |  | Reference |  | Reference |  | Reference |  |  |  |
| 6 to 10 years | 1.59 (0.76 to 3.36) | 0.22 | 1.58 (0.74 to 3.38) | 0.24 | 0.49 (0.18 to 1.39) | 0.18 | 0.82 (0.24 to 2.77) | 0.75 |  |  |
| More than 10 years | 2.08 (1.14 to 3.81) | **0.02** | 1.62 (0.81 to 3.22) | 0.17 | 1.28 (0.65 to 2.51) | 0.47 | 1.93 (0.74 to 5.03) | 0.18 |  |  |
| Born in Australia | 1.63 (0.64 to 4.16) | 0.31 | 1.34 (0.5 to 3.59) | 0.56 | 2.77 (0.97 to 7.91) | 0.06 | 4.28 (1.16 to 15.75) | 0.03 |  |  |
| **Language**** |  |  |  |  |  |  |  |  | **<0.001** | **<0.001** |
| **Information source^^^^** |  |  |  |  |  |  |  |  |  |  |
| Official Australian source/public broadcaster | 0.79 (0.53 to 1.18) | 0.24 | 1.18 (0.71 to 1.97) | 0.52 | 0.76 (0.44 to 1.31) | 0.32 | 1.18 (0.65 to 2.14) | 0.59 | 0.40 | 0.77 |
| Australian commercial source | 0.55 (0.37 to 0.82) | **<0.001** | 0.63 (0.40 to 1.01) | 0.06 | 0.42 (0.25 to 0.71) | **<0.001** | 0.47 (0.26 to 0.84) | **0.01** | **0.001** | **0.02** |
| Social media | 1.03 (0.69 to 1.53) | 0.89 | 1.29 (0.81 to 2.07) | 0.29 | 0.61 (0.36 to 1.06) | 0.08 | 0.48 (0.25 to 0.93) | **0.03** | 0.17 | **0.03** |
| Friends or family living in Australia | 1.58 (1.05 to 2.37) | 0.03 | 1.25 (0.77 to 2.03) | 0.37 | 1.23 (0.68 to 2.21) | 0.50 | 1.07 (0.57 to 1.98) | 0.84 | 0.09 | 0.67 |
| Community | 1.38 (0.92 to 2.09) | 0.12 | 1.09 (0.62 to 1.9) | 0.77 | 0.71 (0.42 to 1.22) | 0.22 | 0.75 (0.39 to 1.45) | 0.40 | 0.06 | 0.59 |
| Overseas information source | 1.52 (1.02 to 2.26) | **0.04** | 0.64 (0.34 to 1.23) | 0.18 | 0.57 (0.33 to 0.99) | **0.05** | 0.39 (0.17 to 0.89) | **0.03** | **0.004** | 0.07 |
| **Comorbid health conditions^^^** |  |  |  |  |  |  |  |  | **0.01** | 0.11 |
| 0 | Reference |  | Reference |  | Reference |  | Reference |  |  |  |
| 1 | 1.62 (1.03 to 2.57) | **0.04** | 1.6 (0.89 to 2.88) | 0.12 | 0.74 (0.39 to 1.4) | 0.35 | 0.80 (0.38 to 1.7) | 0.56 |  |  |
| 2 or more | 1.93 (1.15 to 3.26) | **0.01** | 1.61 (0.84 to 3.06) | 0.15 | 1.64 (0.9 to 2.98) | 0.10 | 1.88 (0.92 to 3.87) | 0.09 |  |  |
| **IRSAD decile**^†^ | 1.06 (0.99 to 1.13) | 0.13 | 1.04 (0.94 to 1.15) | 0.50 | 0.94 (0.85 to 1.04) | 0.23 | 1.01 (0.9 to 1.13) | 0.91 | 0.09 | 0.50 |

*Refence group = ‘yes’ (i.e. willing to get a vaccine); analysis controls for IRSAD and date of survey completion; 1 respondent indicated ‘other/prefer not to say’ and is not included in weighted analysis; **individual comparisons for language group not presented; **^^^**Information sources entered as separate variables as participants could select more than one; ^^^**^^^** Comorbid health conditions included respiratory disease, asthma, chronic obstructive pulmonary disease, high blood pressure, cancer, heart disease, stroke, diabetes, depression or anxiety; ^†^ p value for test of model effect includes multinomial regression (both ‘not sure’ and ‘no’ responses to vaccine intention) ; ^†^Index of Relative Socio-economic Advantage and Disadvantage

**Table S6. Barriers to vaccine willingness***

| **Barrier** | **N** | **%** |
| --- | --- | --- |
| I am worried about side effects | 102 | 30.4 |
| I do not think the vaccine will be safe | 80 | 23.9 |
| I need more information to make a decision | 38 | 11.4 |
| I am worried about how the vaccine may affect my other illness | 24 | 7.1 |
| I want to wait so that i can see how other countries go first | 22 | 6.5 |
| I think the vaccine may not work well | 16 | 4.8 |
| I do not trust the drug companies | 16 | 4.7 |
| Other (E.g. concerns about breastfeeding or interaction with other medicines / other health conditions, not wanting AstraZeneca, feeling their immune system is healthy) | 13 | 3.9 |
| I do not trust the government | 9 | 2.8 |
| I’ve had a bad experience with vaccines in the past | 8 | 2.4 |
| I don’t like needles | 7 | 2.1 |
| **Total** | **335** | **100.0** |

*1 respondent indicated ‘other/prefer not to say’ and is not included in weighted analysis; only one barrier could be selected.

**Table S7. Barriers to COVID-19 vaccine willingness, by language group***

| **Barriers** | **Arabic** | | **Assyrian** | | **Croatian** | | **Dari** | | **Dinka** | | **Hindi** | | **Khmer** | | **Chinese** | | **Samoan/ Tongan** | | **Spanish^^^** | |
| --- | --- | --- | --- | --- | --- | --- | --- | --- | --- | --- | --- | --- | --- | --- | --- | --- | --- | --- | --- | --- |
|  | **n** | **%** | **n** | **%** | **n** | **%** | **n** | **%** | **n** | **%** | **n** | **%** | **n** | **%** | **n** | **%** | **n** | **%** | **n** | **%** |
| I am worried about side effects | 14 | 46.8 | 22 | 28.5 | 18 | 26.6 | 7 | 25.8 | 10 | 32.3 | 4 | 38.3 | 0 | 0.0 | 18 | 44.2 | 4 | 14.5 | 4 | 24.0 |
| I do not think the vaccine will be safe | 2 | 7.6 | 16 | 20.1 | 41 | 60.2 | 3 | 8.9 | 3 | 10.7 | 0 | 0.0 | 0 | 0.0 | 12 | 28.6 | 4 | 11.9 | 0 | 2.6 |
| I need more information to make a decision | 2 | 8.6 | 4 | 4.9 | 1 | 1.4 | 3 | 11.4 | 5 | 16.8 | 6 | 61.7 | 0 | 0.0 | 3 | 8.2 | 11 | 36.2 | 2 | 12.3 |
| I am worried about how the vaccine may affect my other illness | 0 | 1.6 | 8 | 9.7 | 2 | 3.3 | 3 | 12.1 | 1 | 3.3 | 0 | 0.0 | 0 | 0.0 | 2 | 4.6 | 4 | 11.9 | 4 | 25.0 |
| I want to wait so that i can see how other countries go first | 0 | 1.6 | 2 | 2.5 | 0 | 0.0 | 11 | 38.4 | 2 | 6.7 | 0 | 0.0 | 1 | 100.0 | 0 | 0.0 | 1 | 4.3 | 4 | 23.5 |
| I think the vaccine may not work well | 3 | 10.3 | 6 | 7.9 | 2 | 3.3 | 0 | 1.7 | 1 | 3.4 | 0 | 0.0 | 0 | 0.0 | 3 | 6.5 | 0 | 0.0 | 0 | 1.5 |
| I do not trust the drug companies | 0 | 0.0 | 11 | 13.7 | 1 | 1.8 | 0 | 0.0 | 3 | 8.7 | 0 | 0.0 | 0 | 0.0 | 0 | 0.0 | 0 | 0.0 | 1 | 5.3 |
| Other | 5 | 18.2 | 1 | 1.9 | 0 | 0.0 | 0 | 0.0 | 4 | 11.4 | 0 | 0.0 | 0 | 0.0 | 0 | 1.1 | 0 | 0.0 | 1 | 4.2 |
| I do not trust the government | 2 | 5.4 | 4 | 5.5 | 1 | 2.2 | 0 | 0.0 | 0 | 0.0 | 0 | 0.0 | 0 | 0.0 | 0 | 0.0 | 2 | 5.6 | 0 | 1.5 |
| I've had a bad experience with vaccines in the past | 0 | 0.0 | 4 | 4.5 | 1 | 1.2 | 0 | 1.7 | 1 | 2.7 | 0 | 0.0 | 0 | 0.0 | 1 | 1.6 | 2 | 5.6 | 0 | 0.0 |
| I don't like needles | 0 | 0.0 | 1 | 0.9 | 0 | 0.0 | 0 | 0.0 | 1 | 4.0 | 0 | 0.0 | 0 | 0.0 | 2 | 5.1 | 3 | 9.9 | 0 | 0.0 |
| **Total** | **29** | **100.0** | **79** | **100.0** | **68** | **100.0** | **29** | **100.0** | **32** | **100.0** | **9** | **100.0** | **1** | **100.0** | **40** | **100.0** | **30** | **100.0** | **17** | **100.0** |

*1 respondent indicated ‘other/prefer not to say’ and is not included in weighted analysis; only one barrier could be selected. ^^^Spanish language group had substantial gaps in recruitment across age groups
